# The Coviral Portal: Multi-Cohort Viral Loads and Antigen-Test Virtual Trials for COVID-19

**DOI:** 10.1101/2023.05.05.23289582

**Authors:** Alexandra Morgan, Elisa Contreras, Michie Yasuda, Sanjucta Dutta, Donald Hamel, Tarini Shankar, Diane Balallo, Stefan Riedel, James E. Kirby, Phyllis J. Kanki, Ramy Arnaout

## Abstract

**Background:** Regulatory approval of new over-the-counter tests for infectious agents such as SARS-CoV-2 has historically required that clinical trials include diverse groups of specific patient populations, making the approval process slow and expensive. Showing that populations do not differ in their viral loads—the key factor determining test performance—could expedite the evaluation of new tests.

**Methods:** 46,726 RT-qPCR-positive SARS-CoV-2 viral loads were annotated with patient demographics and health status. Real-world performance of two commercially available antigen tests was evaluated over a wide range of viral loads. An open-access web portal was created allowing comparisons of viral-load distributions across patient groups and application of antigen-test performance characteristics to patient distributions to predict antigen-test performance on these groups.

**Findings:** In several cases distributions were surprisingly similar where a difference was expected (e.g. smokers vs. non-smokers); in other cases there was a difference that was the opposite direction from expectations (e.g. higher in patients who identified as White vs. Black). Sensitivity and specificity of antigen tests for detecting contagiousness were similar across most groups. The portal is at https://arnaoutlab.org/coviral/.

**Conclusions:** In silico analyses of large-scale, real-world clinical data repositories can serve as a timely evidence-based proxy for dedicated trials of antigen tests for specific populations. Free availability of richly annotated data facilitates large-scale hypothesis generation and testing.

**Funding:** Funded by the Reagan-Udall Foundation for the FDA (RA and JEK) and via a Novel Therapeutics Delivery Grant from the Massachusetts Life Sciences Center (JEK).

## Introduction

Diagnosis of new infectious pathogens such as SARS-CoV-2 requires development of new diagnostic tests, which must be evaluated and approved by regulatory agencies before they can be used for patient care. Such tests include over-the-counter (OTC) antigen tests, which have been widely used for at-home testing in the context of COVID-19. In order to be approved, a new test must demonstrate a minimum level of clinical performance. Performance is typically measured as the test’s sensitivity, defined as the proportion of true-positive samples that have a positive result, and its specificity, defined as the proportion of true negatives that have a negative result. Clinical performance must be demonstrated in a defined patient population or group and clinical context, for example inpatients as opposed to outpatients. However, at the start of an outbreak, epidemic, or pandemic, there may not be enough information to know whether a test can be expected to perform differently in some patient groups vs. others. Therefore at the start of development, a new diagnostic test may be approved based on its performance in the general population, not specific groups.

In contrast, as time goes on, evidence for clinical differences among specific groups may emerge. As this happens, it becomes reasonable to ask whether a test might perform differently in specific groups, with important implications for how and potentially even whether that test should be used in a given clinical scenario. Ideally, this question would be answered by conducting dedicated trials of the new diagnostic test in each group of interest. Unfortunately, trials are expensive and slow. Also, the number of specific groups that may be of interest is large, since specific subgroups can be defined not only based on demographics (such as age, race, and gender), comorbidities (such as diabetes, heart disease, or immunosuppression), and care settings (inpatient vs. outpatient vs. emergency room) but also by the many possible combinations of each of these characteristics, which is an essential component of precision medicine.^1^ As a result, in practice it is prohibitively difficult to perform many separate trials on specific groups for even a single diagnostic test, much less for the many tests that are likely to be developed in response to a large-scale outbreak, such as has happened in response to the COVID-19 pandemic. This is a problem for regulators, clinicians, and patients alike.

One solution is to apply a new test’s various performance characteristics to real-world data collected in the course of patient care. Such characteristics include results of existing trials as well as analytical (i.e. pre-clinical) operating parameters such as the limit of detection (LOD). The LOD is defined as the lowest concentration of virus that the test can detect in 95% of replicates. It is routinely determined by manufacturers and validated by clinical laboratories before a test is put to use clinically.^2^ The relationship between concentration and detection is usually understood to follow an S-shaped curve;^3^ fitting it requires at least one additional datapoint besides the LOD. The concentration of the virus may be measured as the viral load, most often defined as the number of copies of viral mRNA per milliliter of testing material (copies/mL).

“Real-world data” means the viral-load result of a reference diagnostic test that has already been approved for the general population. Because this data is from the general population, it will presumably include results on many specific patient groups. One can apply the performance of the new test as described above to the set or “distribution” of viral loads from a group to predict what proportion of patients in the group would have tested positive with the new test. This proportion is the sensitivity of the new test for that group. In this way, one can estimate clinical sensitivity without needing a dedicated trial on that group.

In this study, we apply this approach to COVID-19. We use the 46,726 positive SARS-CoV-2 RT-qPCR results our institution has performed as of this writing and use our electronic health record to annotate each result according to the patient’s demographics, comorbidities, and so on. Importantly, we convert each PCR result from a Ct value to a viral load using robust (100% code-coverage) and accurate publicly available software, as previously described.^2, 4–6^ Although PCR results are typically reported simply as positive or negative, qPCR is intrinsically quantitative (the “q” in “qPCR”); we make use of this quantitative information in its natural unit of measure (viral load). This is in contrast to Ct values, which are less useful because they vary inversely with viral load and correspond to different viral loads on different PCR testing platforms.

We focus especially on sensitivity and specificity for infectivity or contagiousness. Contagiousness is of special interest given the public health focus on using quick, inexpensive tests to curtail community transmission in a pandemic.^7, 8^ Treating contagiousness as a function of viral load, contagiousness can be estimated using a virus culture assay we previously described in which a positive patient sample is applied to susceptible cells and monitored for virus replication. After an initial adsorption period, the cells are washed free of the initial virus to eliminate carryover. The supernatant is then sampled on a timescale of days and tested by PCR for the presence of new virus.^9, 10^ The lowest concentration of virus in a patient sample from which new virus can be recovered is the contagiousness threshold. Because cells in culture have no physical or distance barriers, no mucociliary elevator, and no protection via medications or an immune system, we consider this threshold a conservative estimate. We previously demonstrated this threshold is approximately 50,000 copies/mL and has been fairly stable even as the SARS-CoV-2 virus has evolved.^9, 10^

## Materials and Methods

### Institutional review

Institutional Review Board approval was obtained for all described work under Beth Israel Lahey Health (BILH) IRBs 2022P000328 and 2022P000288. The Harvard T. H. Chan School of Public Health IRB20-1979 provided non-human subjects research determination for virus culture work.

### Defining specific patient groups

We extracted the following information from our hospital’s clinical-research data repository: demographics (age, gender, and self-reported race/ethnicity), socioeconomic status (using the median neighborhood household income for the patient’s ZIP code, obtained via the 2020 U.S. census, as a proxy), care setting (inpatient, outpatient, emergency ward, or other institution), presentation/disposition (based on vital signs, which we combined into a measure of initial presentation), outcome (survived, died with COVID-19 as the cause of death, died with COVID-19 as an incidental finding), vaccination status (vaccinated, unvaccinated, or unknown), treatment (CPT-encoded procedures, remdesivir (GS-5734; Gilead Sciences, Foster City, CA) administration, steroid administration), comorbidities (according to the Charlson Comorbidity Index:^11^ body-mass index, diabetes, chronic heart disease, chronic lung disease, chronic renal disease, liver disease, dementia, chronic neurological conditions, connective-tissue disease, human immunodeficiency virus (HIV), and malignancy), and immunosuppression status^12^ (CD4+ T-count <100 cells/µL, hematologic malignancy, chemo/immuno-modulating agent alone or in setting of solid malignancy, organ transplant, or rheumatologic/inflammatory condition). The rationale for extracting these data items specifically was twofold: first, this list includes the complete COVID-19 core diagnostic data at federal and state levels; second, it includes data necessary for calculating the well validated 4C Mortality Score for SARS-CoV-2.^13^ ICD-10 codes corresponding to the listed comorbidities were determined by a physician (Dr. Arnaout) following prior methodologies^14^ but updated for 2022-2023.

At presentation, patients were considered sick if any of the following were true within 1 day of the PCR test sample: systolic blood pressure <90 mmHg, diastolic blood pressure <60 mmHg, heart rate >100 beats per minute, respiratory rate >18 breaths per minute, or temperature >99.1°F. They were otherwise considered well, with the exception that if no values were recorded (NULL in the data repository) for all criteria, presentation was considered unknown and therefore not assigned.

Patients were designated as immunocompromised at the time of PCR testing if one of the following were true: on their most recent T-cell subset analysis report, their absolute CD4+ cell count was <100 cells/µl; they had a diagnosis of either lymphoma or leukemia associated with a healthcare encounter (visit, admission, or phone call) either before the PCR test or within 60 days after the PCR test; they were on any of the following medications on an ongoing basis, prescribed prior to the PCR test and with enough refills to include the time up to 30 days prior to the PCR test: abatacept, adalimumab, anakinra, azathioprine, basiliximab, budesonide, certolizumab, cyclosporine, daclizumab, dexamethasone, everolimus, etanercept, golimumab, infliximab, ixekizumab, leflunomide, lenalidomide, methotrexate, mycophenolate, natalizumab, pomalidomide, prednisone, rituximab, secukinumab, serolimus, tacrolimus, tocilizumab, tofacitinib, ustekinumab, or vedolizumab. Otherwise, they were designated not immunocompromised.

Supplementary Table 1 provides further details for the above methods.

### Viral load

The SARS-CoV-2 RT-qPCR testing in this study was performed on three Abbott Molecular platforms: m2000, Alinity m, and Alinity 4-Plex (Abbott Molecular, Des Plaines, IL, U.S.A.). These detect identical SARS-CoV-2 N and RdRp gene targets. They are extremely sensitive, with LOD of ∼100 copies/mL. They output a quantitative fractional cycle number (FCN), a type of Ct value described in detail elsewhere.^15^ Together these platforms accounted for 46,726 positive tests.

Ct values were converted to viral loads in units of copies of viral mRNA per mL using the public Python package *ct2vl* as previously reported.^4^ Briefly, this software was validated via calibration curves established for all platforms using an extended SeraCare panel (LGC Seracare, Milford, MA) panel based on a SARS-CoV-2 genome incorporated into replication-incompetent, enveloped Sindbis virus and calibrated based on digital PCR at US National Institutes of Standards and Technology (NIST) and LGC/Seracare.^16^ Validation material ranged in viral load from 300 to 10^6^ viral genome copies/mL. Results were harmonized with the cycle threshold for a spiked internal control also amplified in each SARS-CoV-2 assay to confirm lack of PCR inhibition and accurate viral load output. The standards, modeling SARS-CoV-2 virus, were run through all stages of sample preparation and extraction to allow appropriate comparison with identically processed patient samples. *R*^2^ was ∼0.99 for all calibration determinations, indicating assays are robustly quantitative.

### Presumed SARS CoV-2 variant

Presumed variant was inferred from the date of sample collection based on the data presented by Covariants (https://covariants.org) showing the frequency of sequencing particular variants in Massachusetts, the United States, and other locations.^17^ Specimens from before June 7, 2021 were annotated as being an early variant. Specimens from between July 7, 2021 and December 6, 2021 were annotated delta variant. Specimens from after January 3, 2022 were annotated as omicron variant. Results from the month between windows, when more than one major variant was common, were not annotated with a presumed variant and are omitted from by-variant comparisons.

### Evaluation of antigen tests vs. PCR

Patients seeking COVID testing at a drive-through testing site near Boston affiliated with our medical center^5, 6^ between May 23 and November 4 of 2022 were offered the opportunity to participate in a separate arm, providing a comparative, parallel prospective study. This represented community testing for both symptomatic and asymptomatic individuals with diverse demographics (age, race, sex, socio-economic status).

Each patient who consented had both a standard-of-care PCR test and two OTC antigen tests performed (Abbott BinaxNow COVID-19 Ag Card and CareStart COVID-19 Antigen Home Test). The PCR test was performed on material collected with a nasopharyngeal swab. SARS-CoV-2 RT-qPCR testing was performed using the Abbott m2000 RealTime or Alinity m SARS-CoV-2 assays according to the manufacturer’s instructions, yielding, for each positive sample, a Ct value which was converted to viral load as previously described. Specimens for the antigen tests were collected with separate nasal swabs for each test, according to the manufacturer’s instructions. These were collected and the tests performed by study personnel after informed consent was obtained on-site within the time-frame constraints detailed in each test’s instructions for use, as per IRB. In order to extrapolate antigen-test performance from this subset to all patients, positivity vs. viral load was modeled by logistic regression (the LogisticRegression function in Python’s scikit-learn library).^18^ LogisticRegression converges on optimal parameters in a model predicting the probability of a positive test based on viral load. Parameters were predicted separately for each test. The equation for probability was a standard sigmoid constrained to the range 0-1 (i.e., the lowest probability is zero and the highest probability is 1): *p*(test success) = 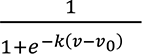 where *v*, the independent variable, is log10 of the viral load. This constraint leaves two free parameters: v0 is the midpoint, i.e. the model’s estimate of where the success rate passes 50%, while *k* controls the steepness, i.e. the change in viral load to change in probability of being positive.

### Contagiousness

Samples were then stored at 4°C until contagiousness testing. This was done within a four-day time period. We previously validated that freeze-thaw does not impact viral viability and will bank remaining samples for future investigations. Quantitative viral culture was performed on a random sample of PCR-positive samples. Vero E6 cells (ATCC CRL-1586) were seeded on a 6-well flat bottom plate at 0.3×10^6^ cells per well in Eagle’s minimum essential media (EMEM) containing 1% antibiotic-antimycotic, 1% HEPES and 5% fetal calf serum (FCS, Gibco) grown to confluence at approximately 1 × 10^6^ cells per well, inoculated with 250µL of patient sample, and incubated at 37°C for 24 hours for viral adsorption, as previously described.^10, 19–21^ Carryover of non-viable viral RNA present in samples was limited by washing cell cultures after the 24-hour viral adsorption and adding fresh EMEM composite media with reduced FCS to 2% for viral growth, meaning detectable virus represents viable replicating virus. On days 3 and 6, cell culture supernatant was removed and added to 800µL of VXL buffer (QIAGEN, German, MD) (1:1 ratio) for subsequent nucleic acid extraction and detection of virus by PCR. Viral load in culture supernatants on days 3 and 6 served as a quantitative surrogate for viable (i.e. replication-competent) virus in the patient sample and provided a measure of the magnitude of sample infectivity. SARS-CoV-2 RT-qPCR testing of Vero cell culture supernatants was performed using the Abbott m2000 Real-Time or Alinity m SARS-CoV-2 assays according to the manufacturer’s instructions. The contagiousness threshold was determined by the threshold patient-sample viral load value resulting in detectable culture viral load.

### Whole-genome viral NGS

Next-generation-sequencing (NGS)-based sequencing of select PCR-positive samples from the viral antigen evaluation study was performed as follows. Full-length SARS-CoV-2 viral genome sequencing was performed on the Oxford Nanopore MinION system (≥R9.4 flowcell; Oxford Nanopore Technologies-ONT, Oxford, UK) using the guppy basecaller and the downstream ARTIC network bioinformatics pipeline for genome assembly.^22, 23^ The workflow was run on a 2021 Intel Core i9-11900 Rocket Lake 3.5GHz 8-core LGA 1200 boxed processor with NVIDIA A5000 GPU. Standard coverage and quality metrics and plots were produced, single-nucleotide variants were recorded, and variants assigned using NextClade.^24^

### Web portal and privacy protection

The portal was written using Svelte and d3 for the interactive frontend and Python run against a Postgres database for the backend. To reduce re-identification risk, ages were jittered by adjusting the patient’s date of birth by a random number of days (drawn from a Gaussian distribution with a standard deviation of two years) before calculating patient’s age at the time of each test. Groups smaller than 4-8 patients are suppressed and therefore not viewable. Revealing exact sizes of such small groups defined by multiple patient characteristics would pose a re-identification risk. To prevent inferring the sizes of these groups by subtraction of viewable group sizes, viewable group sizes are jittered by dropping approximately 0.5-1% of the data on any split by patient feature. To maximize consistency of the results of jittering as data are updated, jittering was performed using random number seeds based on pseudo-identifiers (which are never uploaded and thus inaccessible to/safe from the web client). For ease of visualization, plots of viral load distributions are shown as kernel-density estimates (i.e. smoothed) using a Gaussian kernel of width 0.25 log10 viral load units (∼1.7-fold).

### Statistical tests

The geometric mean viral load for each patient group was calculated as a summary statistic. The geometric (as opposed to arithmetic) mean was chosen because viral loads vary over many orders of magnitude.^2^ The Kolmogorov-Smirnov test (KS; scipy.stats.kstest) was used to compare distributions. This test was used because data were not distributed normally and KS does not require normality (unlike, for example, the t-test, which requires normal distributions). KS tests the null hypothesis that the distributions of viral loads for two patient groups are statistically indistinguishable.^25^ The p-value gives the probability that distributions from the two groups are drawn from the same underlying distribution. A large p-value means the two groups are statistically indistinguishable; a small p-value means they are different. Interpretation of p-values as significant vs. not significant requires a significance threshold, which requires correction for multiple comparisons if multiple comparisons are performed.^26, 27^ Because the number of comparisons performed via the web portal is up to the user, un-corrected p-values are reported, with interpretation as significant or not significant left to the user.

### Software and hardware

Data extraction, annotation, statistics, and analyses were performed using standard Unix tools and Python 3.9+ using the pandas, numpy, scipy, and scikit libraries and the interactive Jupyter notebook environment. Figures were created using Python graphics libraries matplotlib and seaborn, and OmniGraffle 7 (The Omni Group, Seattle, WA).

## Results

### A web portal for large-scale real-world SARS-CoV-2 viral load results for different patient groups

46,726 COVID-19 PCR results representing approximately 39,180 unique individuals were converted to viral loads and annotated for patient demographics, comorbidities, presentation, treatment, and socioeconomic status and made available for interactive investigation via a public web portal at https://arnaoutlab.org/coviral/ (Table 1). The portal^28, 29^ allows users to visualize the viral load distribution for any patient group, to compare distributions between groups, and to estimate, for each group, the sensitivity and specificity of a given OTC test for detecting contagious individuals. Users can define and compare complex subgroups by selecting multiple characteristics via checkboxes in the user interface (Fig. 1). In this work, all the figures that contain distributions are direct screenshots from the portal.

**Fig. 1:**
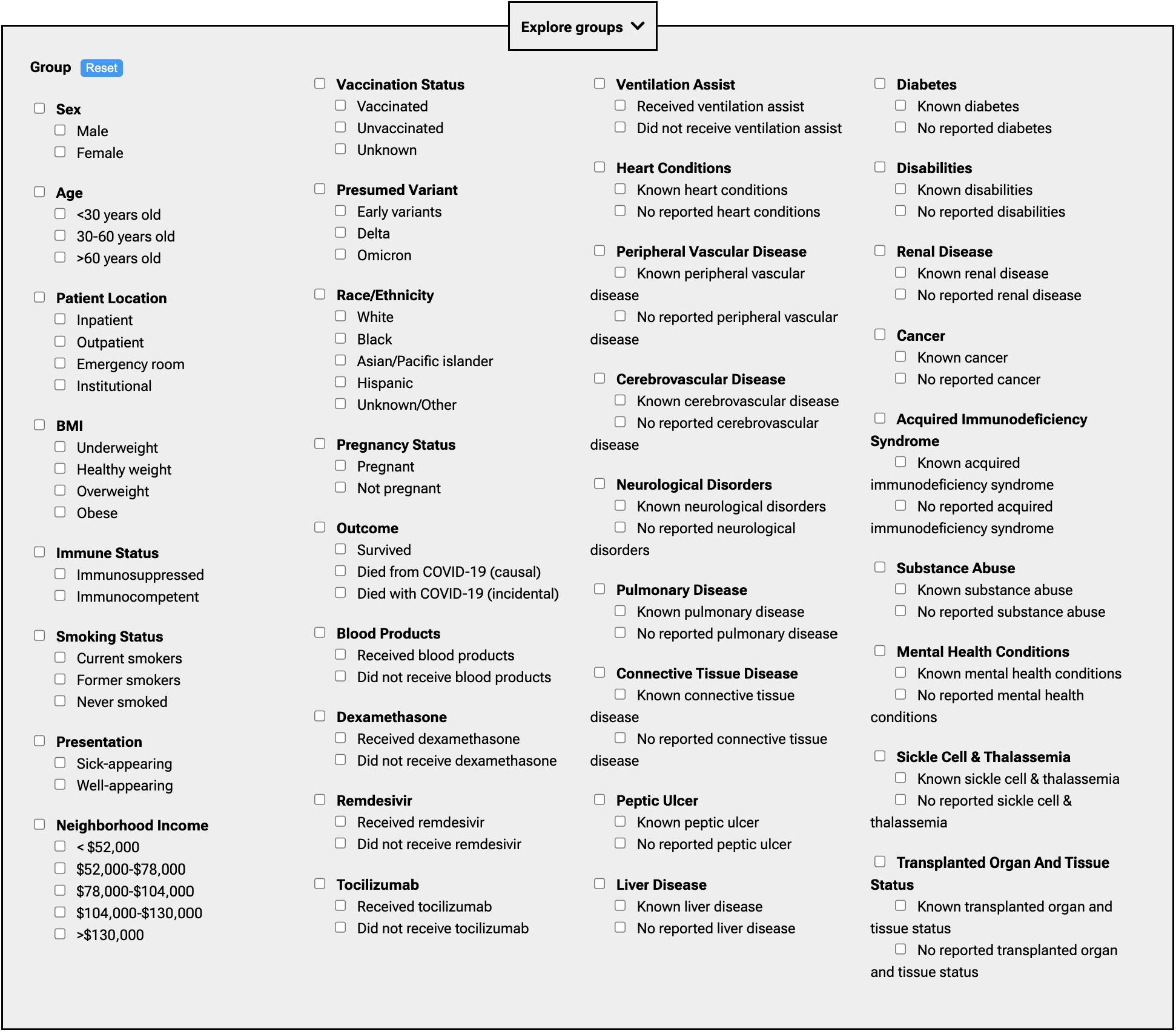
Patient characteristics available for defining patient groups and subgroups. This screenshot from the web portal demonstrates the ability for users to define patient groups by demographic, comorbidity, treatment, vaccination status, pandemic era, and so on, using checkboxes.

**Table 1:** Summary of patient characteristics. Shown are counts for select high-level categories as of April 14, 2023. Counts may differ somewhat from the counts presented through the web portal as a result of jittering and as more data continues to be added through the portal. Note that counts broken down by characteristics do not add up to the total, because of the nulling out of some data to reduce re-identification risk (see Methods).

|  |  |  |
| --- | --- | --- |
| 543 | **Sex** |  |
|  | Female | 25,884 |
| 544 | Male | 20,608 |
|  | **Age** |  |
|  | <30 years old | 12,446 |
| 545 | 30-60 years old | 21,889 |
|  | > 60 years old | 12,100 |
|  | **Self-reported Race or Ethnicity** |  |
| 546 | Unknown/Other | 14,530 |
|  | White | 13,806 |
| 547 | Black | 8,299 |
|  | Hispanic | 7,540 |
| 548 | Asian/Pacific Islander | 2,472 |
|  | **Setting** |  |
|  | Inpatient | 2,157 |
| 549 | Outpatient | 11,758 |
|  | Emergency room | 1,779 |
| 550 | Other institutions | 31,031 |
|  | **Variant** |  |
|  | Early | 28,289 |
| 551 | Delta | 2,911 |
|  | Omicron | 11,264 |
|  | **Vaccination status** |  |
| 552 | Vaccinated | 6,806 |
|  | Unvaccinated | 6,960 |
| 553 | Unknown | 32,732 |
|  | **Outcome** |  |
|  | Died from COVID-19 | 398 |
| 554 | Died with COVID-19 as an incidental finding | 143 |
|  | Survived | 45,938 |
| 555 | **Testing platform** |  |
|  | Abbott m2000 | 24,243 |
|  | Abbott Alinity | 20,593 |
| 556 | Abbott Alinity 4-plex | 1,889 |
|  | **Total** |  |
| 557 |  | 46,726 |

### Overall viral load distributions

Viral loads varied over nearly 10 orders of magnitude, from 7 copies/mL (the lowest our system will report) to 1.5 billion copies/mL (99^th^ percentile). This extraordinary range is consistent with observations from early in the pandemic (spring-summer of 2020).^2^ The referenced early observations suggested that viral loads were, to a good approximation, distributed fairly uniformly over the range. In contrast, the current dataset, which is ten times as large (46,726 results vs. 4,774 in the previous work^2^), demonstrates clear bimodality: patients’ viral loads tend to be either very low, with a peak around the LOD of 100 copies/mL, or else very high, with a peak around 100 million copies/mL (Fig. 2). This bimodality is apparent in retrospect (e.g., Fig. 2a of Arnaout et al. 2021^2^) but required a large dataset to visualize clearly. Further research is needed to understand the reason(s) for these two peaks.

**Fig. 2:**
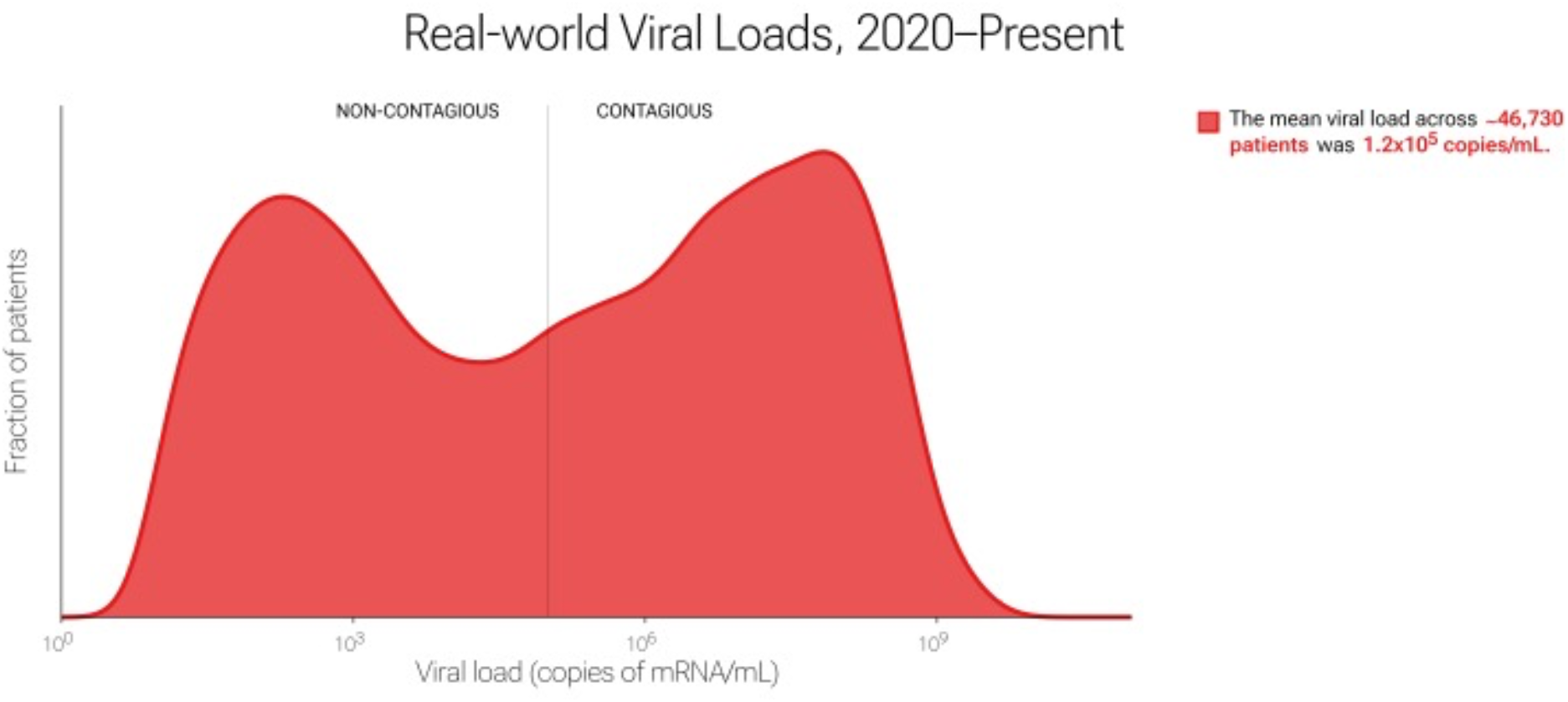
Overall bimodal distribution of viral loads. When no checkboxes are selected to constrain or partition the dataset, users see the distribution of all viral loads. The marked bimodal distribution is clearly apparent.

### Viral load comparisons among patient groups: remdesivir treatment and patient presentation

The web portal allows statistical comparisons of many thousands of specific patient groups. Here we describe several examples that are illustrative of the questions that can be asked and answered using this resource. Remdesivir (Gilead Sciences, Foster City, CA) is an intravenously administered RNA polymerase inhibitor initially approved by the FDA for treatment of SARS-CoV-2 in hospitalized adults and adolescents.^30^ Of the 46,726 test results in our dataset, 688 were from patients who then received remdesivir treatment. In practice, at our institution, remdesivir was used for sicker patients. We hypothesized that viral loads would be higher on average in patients who received remdesivir and in sicker patients. The data supported this hypothesis: viral loads were higher on average in both remdesivir-receiving and sicker-appearing patients, with means of 8.6×10^5^ copies/mL in patients who received remdesivir vs. 1.2×10^5^ in those who did not (Fig. 3a) and 9.4×10^4^ in sick-appearing patients vs. 4.1×10^4^ in well-appearing patients (Fig. 3b). In both cases, the difference was due to a greater fraction patients in the high-viral-load peak. This was especially clear in the remdesivir comparison (Fig. 3a). In each case, the KS p-value was 4.3×10^-16^, which we interpret as rejecting the null hypothesis of no difference, with high confidence. These are examples in which the data confirmed hypotheses regarding differences in viral load.

**Fig. 3:**
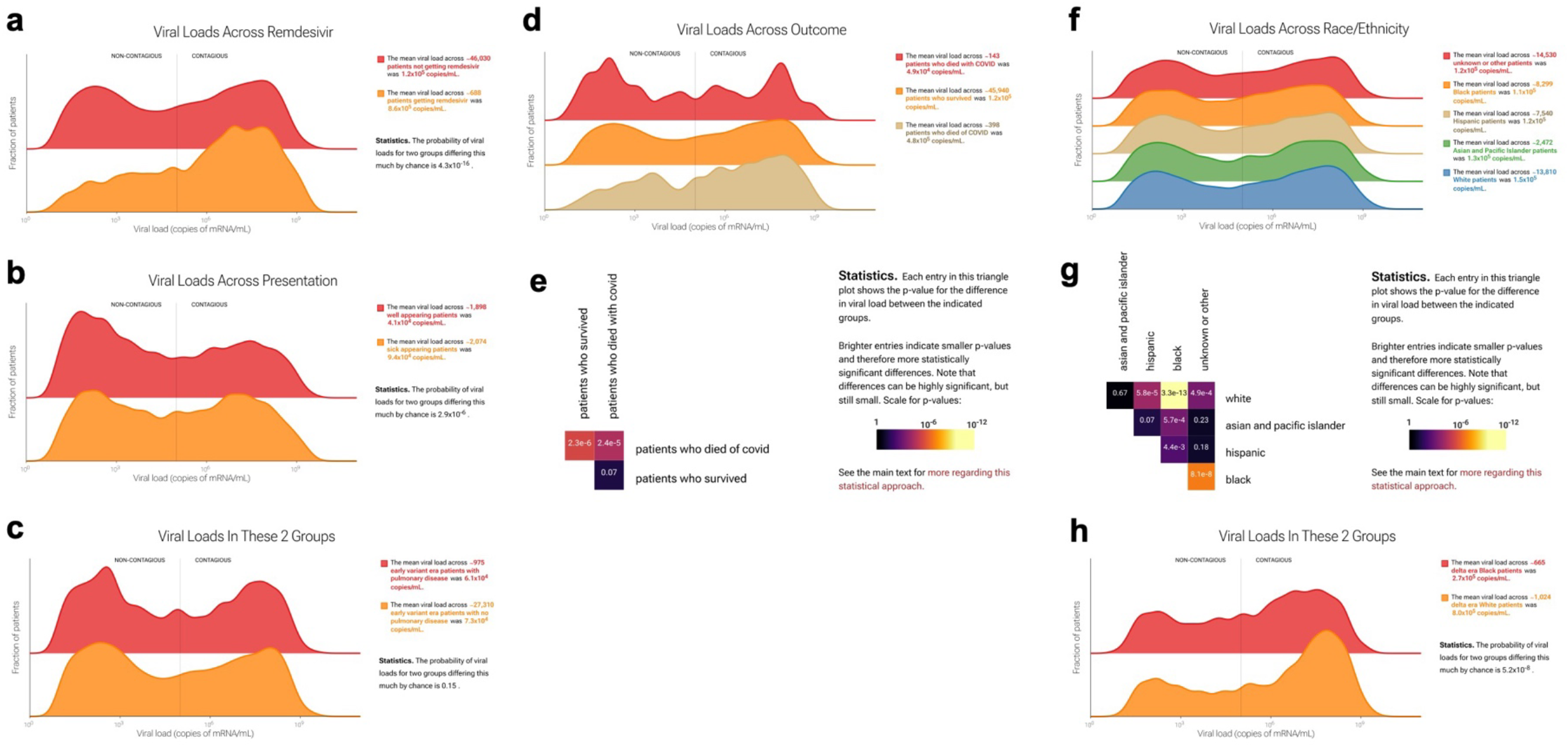
Viral load comparisons by remdesivir treatment, patient presentation, and outcome. Screenshot from the web portal. Viral loads are higher in (a) patients who received remdesivir vs. patients who did not receive remdesivir and in (b) patients who presented as ill-appearing vs. patients who presented as well-appearing (see Table S1 for precise definitions). Note the bimodal distributions, with a low-viral-load peak and a high-viral-load peak. (c) Viral load distributions and pairwise p-values by presence or absence of pulmonary disease for patients during the early variant era. (d) Patients who died from COVID-19 had higher viral loads than either patients who died with COVID-19 as an incidental finding or survivors. Viral loads for were statistically indistinguishable between the latter two groups. The web portal displays distributions in a ridgeline plot from lowest to highest mean, top to bottom. (e) Because there are three or more groups, p-values are displayed as a heatmap, accompanied by explanatory text. Because KS p-values are symmetric, only the top half of the heatmap is shown. Fig. 3f-h: viral load distributions by self-reported race and by race-plus-presumed variant. (f) Viral load distributions by race. (g) Statistical comparisons among these distributions. (h) Viral load distributions between Black vs. White patients in the delta-variant era.

### Unexpected findings: pulmonary disease

Serious cases of COVID-19 are marked by life-threatening respiratory distress. This evolution became less common with the emergence of the omicron strain and the increasing prevalence of prior immunological exposure including vaccination. We hypothesized that patients with pulmonary disease would have higher viral loads than patients without pulmonary disease, especially for early viral variants, which had a stronger tropism for lung as opposed to the upper respiratory tract. However, this hypothesis was not supported (Fig. 3c). Viral loads for the 975 patients with pulmonary disease tested during the early-variant era were statistically indistinguishable from those for the 27,308 patients with no pulmonary disease who were tested during the same time period. This is an example of unexpected findings that this dataset and its web-portal interface can reveal.

### Comparisons among multiple groups: survivorship and causes of death

The web portal also allows users to compare more than two groups of patients at a time. In quantifying mortality during the pandemic, one distinction of value has been between individuals who died with COVID-19 as the proximal cause of death and individuals who died with COVID-19 as an incidental finding. We compared these two groups with survivors (Fig. 3d). We found that the 398 patients who died from COVID-19 in our dataset had higher viral loads than either of the other two groups, and that viral loads were statistically indistinguishable between the approximately 46,000 survivors and the 143 patients who died with COVID-19 as an incidental finding (*p*=0.07). For ease of comparison, the web portal displays distributions in a ridgeline plot from lowest to highest mean, top to bottom. When there are three or more groups, p-values are displayed as a heatmap, accompanied by explanatory text. (Because KS p-values are symmetric, only the top half of the heatmap is shown.)

### Complex patient subgroups: race and presumed variant

The ability to interrogate complex subgroups by checking multiple boxes in the web-portal interface allows more subtle investigations. For example, Black patients have experienced disproportionate morbidity and mortality during the pandemic.^31^ However, the 13,806 patients who self-reported as White in our dataset on average had slightly higher viral loads than the 8,299 who self-reported as Black (KS *p*=3.3×10^-13^). That the viral loads in the White group were on average *higher* suggests that differences in outcome between these groups are not explained by differences in viral load (Fig. 3f-g), despite the clear relationship between viral load and survivorship described above (Fig. 3d). Interestingly, the observed difference is more pronounced during the delta-variant wave (Fig. 3h). During the delta wave (July 7, 2021 to December 6, 2021), viral loads were on average three times as high for White patients (8.0×10^5^ copies/mL, *n*=1,024) as Black patients (2.7×10^5^ copies/mL, *n*=665; KS test *p*=5.2×10^-8^) with a distinctly sharper high-viral-load peak in White patients. This difference was greater in patients over 30 years old and was almost entirely absent in patients under 30 (30-60 y.o: 398 Black patients and 719 White patients, *p*= 2.5×10^-5^; <30 y.o.: 266 Black vs. 299 White patients, *p*=0.14). In contrast, viral load distributions for Black and White patients were more similar for both early in the pandemic and during the omicron variant time period (*p*=4.0×10^-5^ for 4,393 Black and 7,042 White patients and *p*=0.02 for 2,295 Black and 4,341 White patients, respectively). This example illustrates the utility and (statistical) power of the portal for investigating subgroups of interest.

### Antigen test performance

In the head-to-head comparison of PCR and antigen test results, 281 patients consented to participate. Of the PCR samples collected, 277 were tested; the remaining four were mishandled or leaked. Of the 277, 65 had a positive COVID-19 result by PCR (23%). The PCR-positive samples were all tested on either the Alinity m SARS-CoV-2 real time RT-PCR assay or the Alinity m Resp-4-Plex PCR assay. Viral loads in the PCR-positive patients ranged from approximately 10 to approximately 10^9^ copies/mL, with a peak in the distribution between 10^6^ and 10^8^. Of the 65 positive samples, 3 were sequenced and 20 selected at random were used to assess contagiousness in viral culture.

### Antigen test performance

Of the 65 patients with positive PCR tests, 43 tested positive on the Binax antigen test and 40 tested positive on the CareStart antigen test. No invalid antigen tests (lacking the control line) were observed. Only one of the patients who tested negative by PCR tested positive on the antigen tests (both Binax and CareStart), confirming the high specificity of these tests. The proportion of positive antigen tests varied with viral load. At viral loads less than 10^3^ copies/mL, both antigen tests were always negative; at viral loads greater than 10^7^ copies/mL, both antigen tests were always positive. However, there was an overlap of antigen-test-positive and antigen-test-negative results at intermediate viral loads (Fig. 4a). *k* and *v*_0_ values (see Methods) were comparable between the two antigen tests (*k*=1.184, *v*_0_=4.538 for Binax and *k*=1.142, *v_0_*=4.995 for CareStart). The resulting S-shaped curves were used to predict antigen test performance in the web portal.

**Fig. 4:**
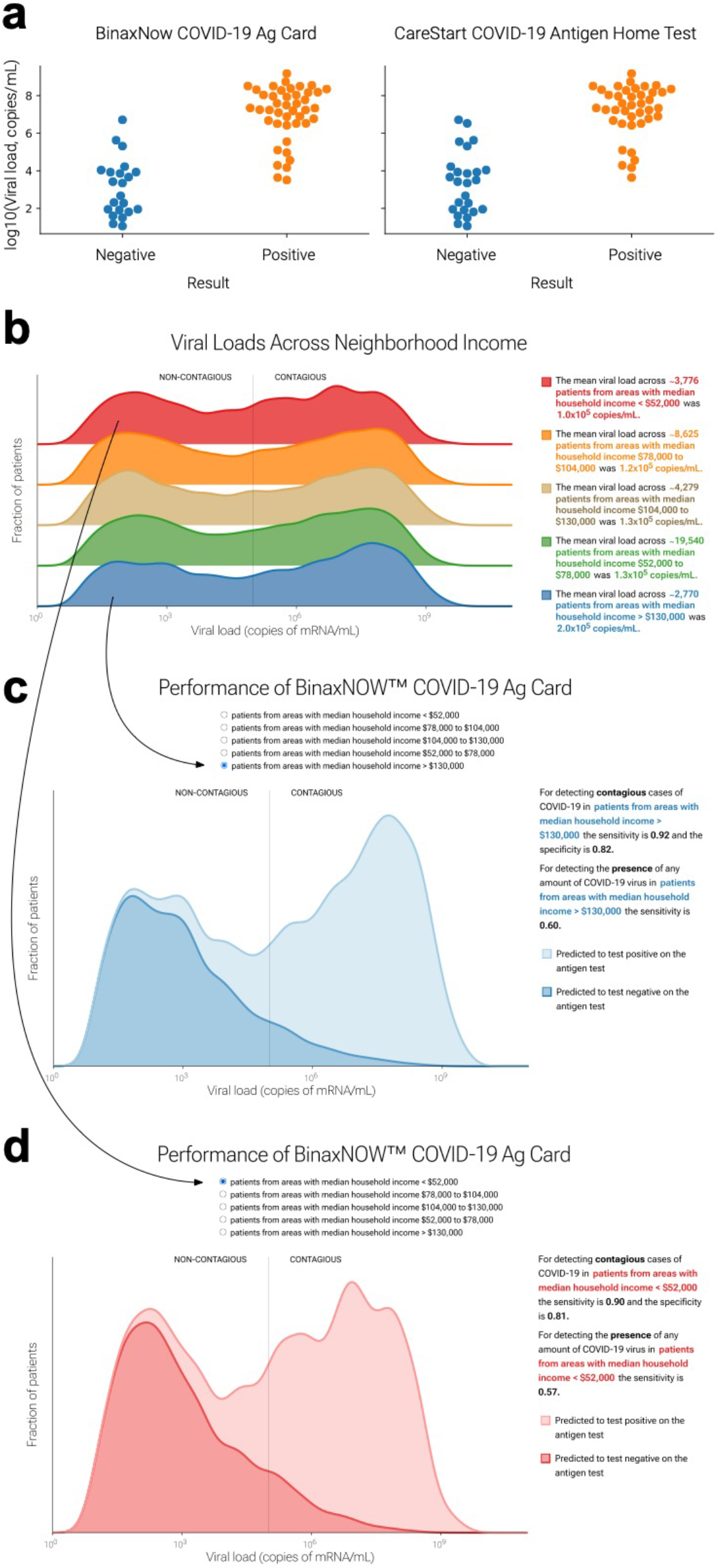
Antigen test results from head-to-head trial and performance on patient subgroups. (a) Each antigen test result for each PCR-positive patient, vs. log_10_ of viral load according to the simultaneous PCR test. (b) Antigen test performance on patients in neighborhoods stratified by median household income. (c) Performance of BinaxNow COVID-19 Ag Card on patients with median household income >$130,000. (d) Performance of BinaxNow COVID-19 Ag Card on patients with median household income <$52,000. The user can use the radio buttons to select any of the patient groups in the viral load distributions in the top section of the web portal. The imputed positive antigen-test results will appear as a lighter shade, and the negative results a darker shade.

The OTC antigen tests that have been widely available on the market since 2021 are considerably less sensitive than RT-qPCR for detecting SARS-CoV-2 infection. However, because their LODs are generally above the contagiousness threshold, they are quite sensitive for detecting contagiousness.^10^ Based on our clinical experience, we hypothesized that antigen tests would perform similarly on different patient groups and subgroups; this hypothesis was largely supported (Fig. 4b-d). The web portal also allows users to estimate sensitivity and specificity for the BinaxNow COVID-19 Ag Card and CareStart COVID-19 Antigen Home Test, based on the modelled performance curves, on any sufficiently large user-selected patient group (Fig 6). The two tests performed well: sensitivities for detecting contagiousness were roughly 0.85-0.90 across patient groups.

### Contagiousness for omicron-era virus

For early-pandemic and delta-wave strains of SARS-CoV-2, the threshold viral load for contagiousness has previously been found to be approximately 10^5^ copies/mL.^10^ For omicron variants, we found that the threshold is statistically indistinguishable from this, at 4.5×10^4^ copies/mL (confidence interval, 1.1×10^4^-1.9×10^5^ copies/mL; *p*=0.23, Fig. 5). The omicron threshold was based on 20 PCR-positive results from our head-to-head clinical trial of 277 total patients. We confirmed that the dominant strain circulating in the Massachusetts Bay area was omicron (BA5.2/Clade 22B) via next-generation sequencing, with only rare single-nucleotide differences relative to those already described (Fig. 6a-d). Because the strain mix in Massachusetts (Fig. 6e) has been highly representative of the strain mix in the country as a whole throughout the pandemic (Fig. 6f), these results support the generalizability of these findings from a particular geographic area to the entire population.

**Fig. 5:**
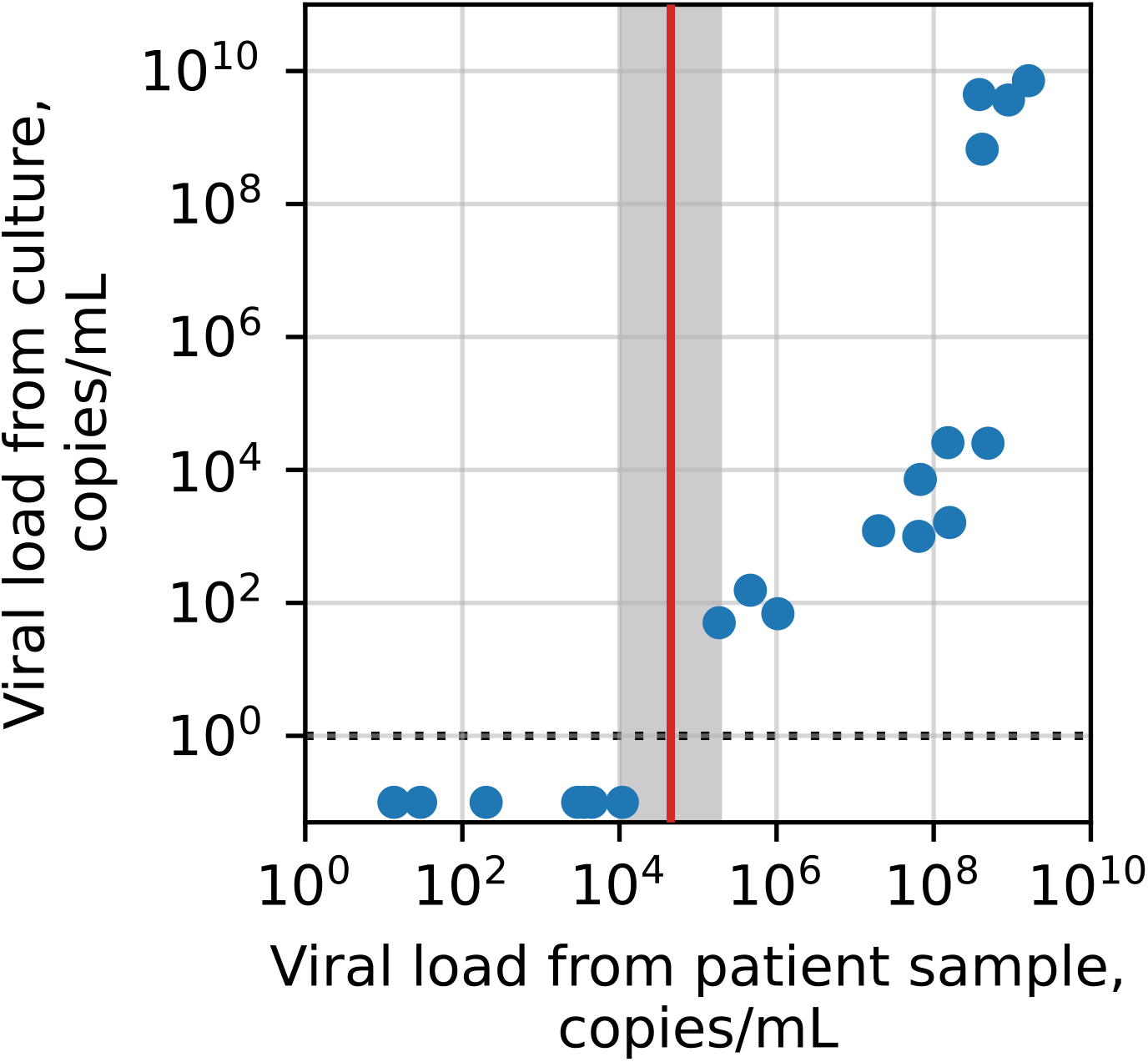
Determination of the contagiousness threshold for omicron-era SARS-CoV-2 strains. Below a certain viral load in the patient sample (x-axis), no virus was recoverable from culture (y-axis, maximum of day 3 and day 6 supernatants). For viewing convenience, culture-negative samples are plotted at 0.1 copies/mL (dotted line, 1 copy/mL). The gray region shows the confidence interval for the threshold. The red line shows the midpoint of this region (on the log10 scale), 45,000 copies/mL, as the most likely threshold.

**Fig. 6.**
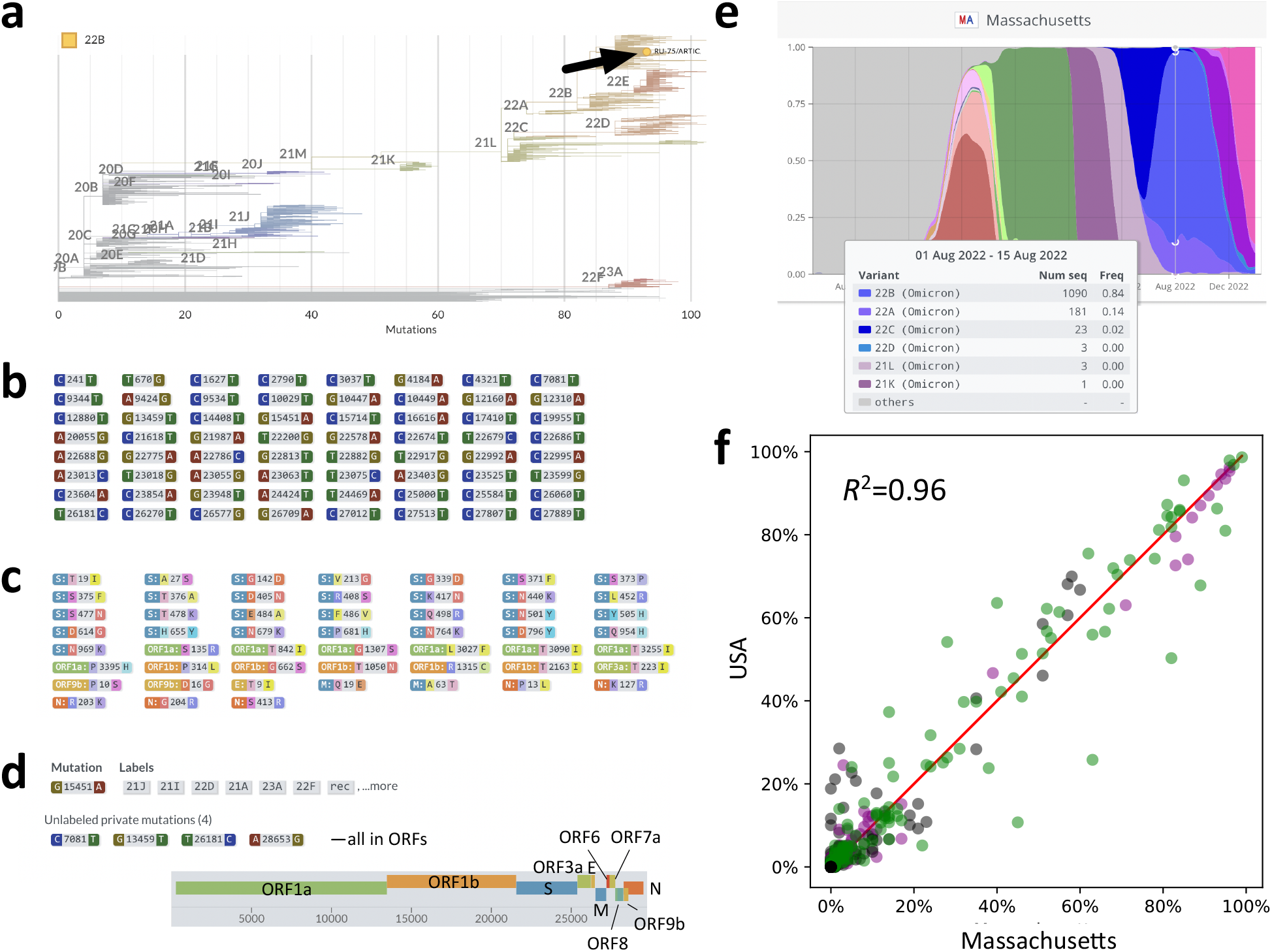
Sequencing late-2022 strain and generalizability of Massachusetts-level results to the United States as a whole. Results of sequencing of a BA5.2/Clade 22B patient sample from Aug.-Sep. 2022 (97.6% coverage). (a) Sample relative to COVID-19 phylogeny (with clade labels). (b) First 64 of the 72 nucleotide substitutions relative to the original Wuhan strain. (c) 52 amino acid substitutions relative to the Wuhan strain. (d) The five unique (“private”) mutations relative to the phylogenetic tree. (e) Distribution of strains in Massachusetts near the time of the sample according to covariants.org. (f) Comparison by frequency of the strains circulating in Massachusetts to those circulating in the United States at the same times demonstrating generalizability of Massachusetts-state variant patterns to the country as a whole. Red line, 1:1. Gray, early strains; purple, delta strains; green, omicron strains. *R*^2^ is for least-squares linear regression of USA vs. Massachusetts data (regression slope=0.97, intercept=0.00).

## Discussion

The COVID-19 pandemic has proven a catalyst for accelerating medical advances, including the development of more efficient methods for developing and testing critical diagnostic assays.^32–37^ It has also drawn attention to the value of reliable public data, including large public datasets.^38–42^ Here we describe such a dataset, to our knowledge the first large dataset of SARS-CoV-2 viral loads in patients across the history of the pandemic through to the present day. The rich clinical annotations of this dataset reveal similarities and differences in viral loads among patients by demographics, presentation, and comorbidity as well as by vaccination status, treatment, and socioeconomic status. Protected patient data was safeguarded by multiple mechanisms. The data revealed cases in which differences were expected as well as cases in which they were unexpected. The size of the dataset and extent of the annotation allow more comparisons than can reasonably be summarized in a single publication; availability of this dataset via the web allows anyone—clinicians, investigators, developers, regulators, and patients, alone or with the assistance of artificial intelligence and/or machine learning (AI/ML)-based tools—to explore and conduct research, to test existing hypotheses, and to generate new research questions.

The clinical utility of measuring and investigating viral load in SARS-CoV-2 has been demonstrated.^43–46^ This is consistent with both the advantage of viral loads relative to Ct values^47, 48^ and their utility in earlier viral infections such as HIV and hepatitis C (HCV). SARS-CoV-2 viral loads have already proven useful in the development and characterization of COVID-19 diagnostics in multiple contexts, including testing on nasopharyngeal secretions, nasal secretions, and saliva.^5,6^ Here we demonstrate their potential to regulators as a tool to streamline evaluation of new OTC tests. Specifically, we demonstrate that a test’s analytical performance measure, namely the LOD, can be used to estimate that test’s clinical sensitivity for detecting contagiousness in any patient group, without having to conduct a dedicated clinical trial for that group. The alternative of innumerable dedicated trials is beyond reasonable expectations of the financial capability of developers or the bandwidth of their clinical testing partners. For this reason regulatory agencies such as the FDA have expressed interest in methods that use large-scale real-world data to streamline test evaluation.

Two assumptions implicit in this approach are worth mention. First, we model the success rate of antigen tests solely as a function of viral load. The assumption is that no other non-negligible factor varies systematically between patient groups. Second, we assume that the curve— success rate as a function of viral load—can be adequately predicted from the data from a fairly small study. There are two possible sources of error in this curve: sampling error, which can be reduced by increasing the number of subjects sampled; and lack-of-fit error, the error inherent in trying to fit a function of the wrong form. The function used in the web portal’s calculations, was chosen on the basis of its history of use in dose-response-type situations, but carries the (reasonable) assumptions that the probability increases smoothly and continuously with increasing viral load, and approaches 0 with sufficiently low viral load and 1 with sufficiently high viral load. These constraints leave only two free parameters, which is desirable for statistical power and robustness to the noise inherent in any such study (e.g., sampling error and measurement error).

A user of the web portal who is accustomed to thinking of test quality solely in terms of LOD (limit of detection) might initially be surprised that the portal prefers two parameters, not one, to define an antigen test’s performance. In effect, the LOD parameter sets the location of the S-shaped curve that relates viral load and performance, and the second parameter—here, the 50% detection threshold—sets the S-shaped curve’s steepness. Without the second parameter, one could fit a curve in which the sensitivity is 0% at all viral loads below the LoD, and 95% at any higher viral load, which is clearly quite different from the relationship observed in the head-to-head study. How different the true shape of the antigen test performance curve is from the logit function used here, and thus whether fitting a different function would work better, can be elucidated by larger head-to-head studies; however, in the midst of a public health emergency, the cost in time of sampling more subjects must be weighed against the value of complete certainty. At any rate, the fitting error was low.

Three important limitations to the dataset in its current form also deserve mention. The first is due to incompleteness of some of the data fields, for example presentation and vaccination status (Table 1). Presentation information was only sometimes available in structured form in our data repository; we did not attempt to extract data from notes to complement incomplete records. Vaccination status was likewise only sometimes available in a structured manner; integration with state-level records could potentially fill in missing records. Second, patient-level annotations are not yet available for download as part of the dataset. Making viral loads freely and easily available for patient groups required significant attention to avoid potential loopholes that might risk patient privacy via identifiability. Methods included suppressing data transmission for groups that were small enough to present potential “journalist risk,”^49^ jittering counts to prevent deduction of the sizes of suppressed groups, and rounding viral loads to two log-scale decimal places.^50^ Further work is necessary to make patient-level annotations available. Third and finally, the size of the dataset, while large, is still insufficient for the smallest groups—for example, cystic fibrosis patients, patients who recently delivered, or Native Americans—to be sufficiently sizeable to draw robust conclusions. One solution is to add data from other care-giving institutions that performed substantial COVID-19 testing; another is to supplement existing large datasets, for example the 50-million-person CVD-COVID-UK initiative, with viral loads. The free availability of methods to convert from Ct values to viral loads facilitates such advances.^4^

As part of the drive toward precision medicine, clinical care benefits from personalization of diagnostic testing: the right test for the right patient, where the importance of patient heterogeneity is increasingly accepted. We have demonstrated that large-scale real-world data can assist this effort by enabling the estimation of personalized clinical sensitivities and specificities without the need for dedicated clinical trials on every patient group. This work is generalizable beyond COVID-19. Laboratory testing is an exceptionally rich source of real-world medical information. It is the highest volume medical activity, with some 20 billion tests performed each year in the United States alone. It is also the most cost-effective, costing just pennies on the healthcare dollar. It is integral to decision-making across medicine, for patients at every level of acuity, from screening to emergencies. Its results are almost always numerical or categorical, making it especially amenable to modern approaches like machine learning. And computational re-analysis is substantially less expensive than de novo trials. The present work supports the view that meaningful value can come economically from additional repurposing of the vast stores of real-world laboratory results that exist in healthcare institutions.

## Data Availability

All data produced are available online at https://arnaoutlab.org/coviral

## Acknowledgements

The authors acknowledge Elliot Hill for assistance on plugging ct2vl into the data processing workflow; to Timothy Graham, Gail Piatkowski, Baevin Feeser, and Griffin Weber for their advice and guidance on extracting data from EMR databases; to funding from the Reagan-Udall Foundation for the FDA (R.A.); and to funding via a Novel Therapeutics Delivery Grant from the Massachusetts Life Sciences Center (J.E.K.). We would also like to acknowledge Abbott (Scarborough, ME) for provision of the Binax Now antigen tests and Ginkgo Bioworks (Boston, MA) for provision of CareStart antigen tests used in this study.

## Author Contributions

Conceptualization, J.K. and R.A.; Software, A.M.; Investigation, D.H., E.C., S.D., and M.Y.; Resources: S.R.; Data Curation, A.M.; Writing — Original Draft, R.A. and A.M.; Writing — Review & Editing, P.K. and J.K.; Visualization, T.S., D.B., A.M., and R.A.; Supervision, R.A; Funding Acquisition, P.K., S.R., and R.A.

---

**Table S1:**
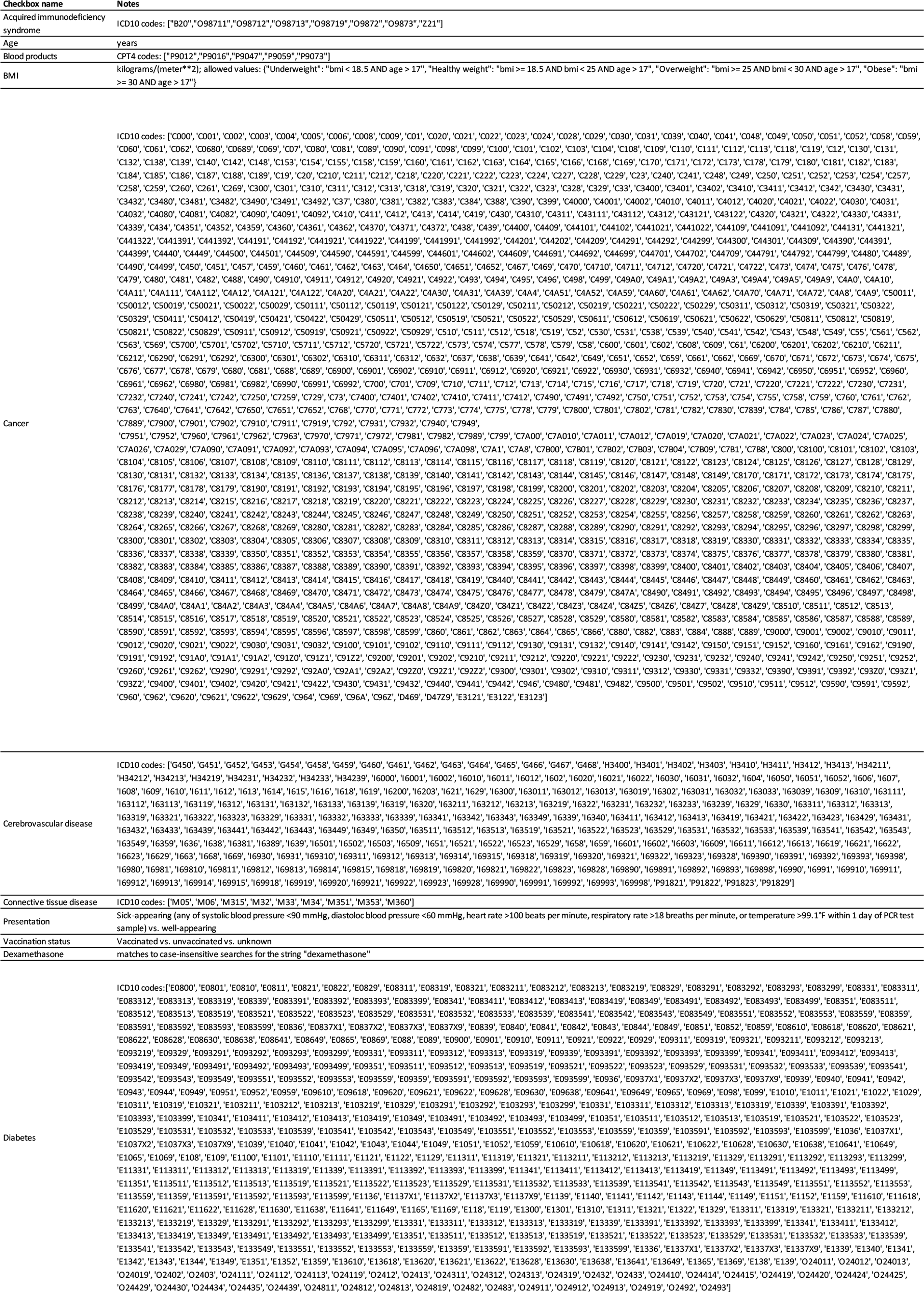
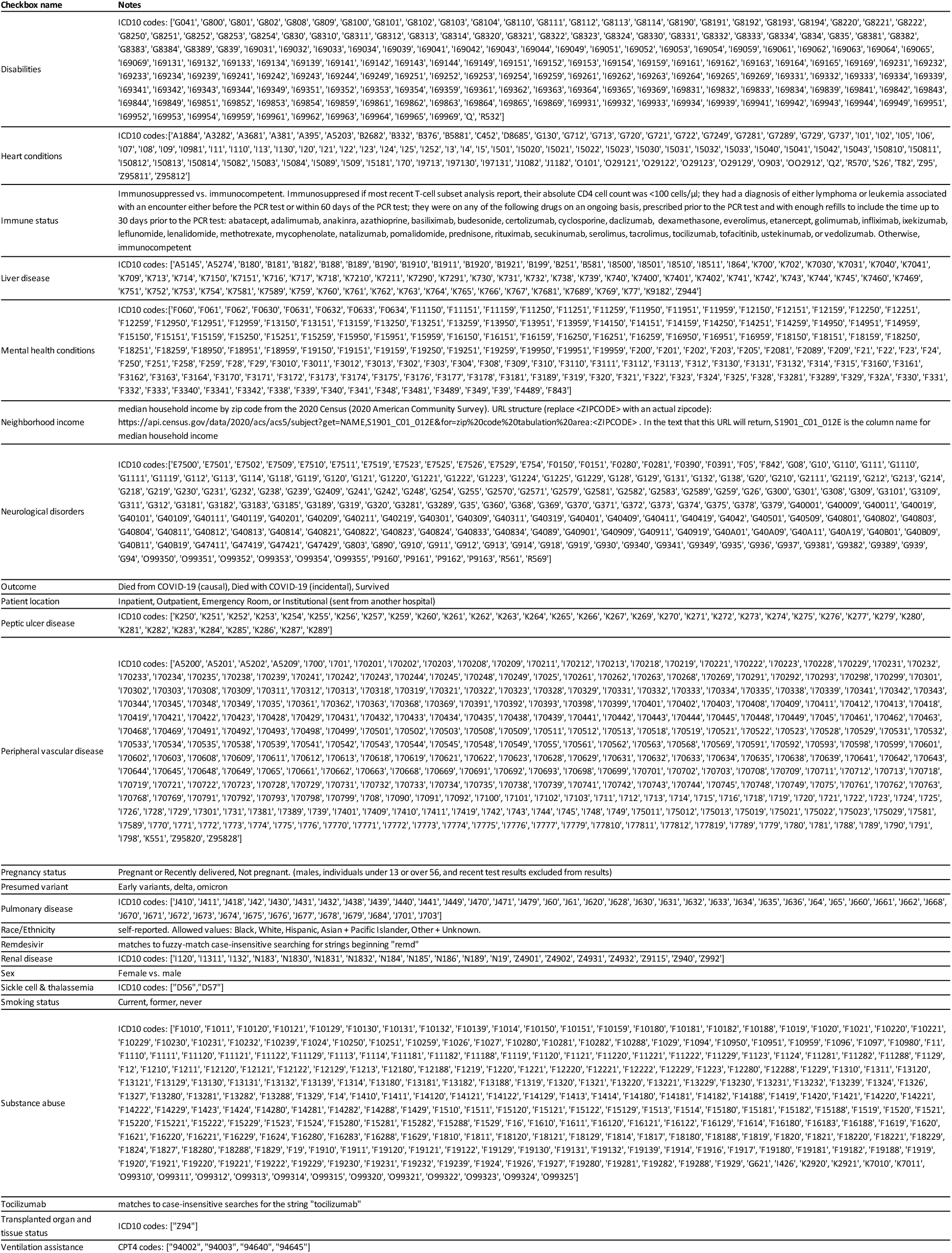
Detailed criteria for defining patient characteristics, Related to Methods. Criteria for each of the characteristics available on the portal are described.

